# Simple, accurate calculation of mechanical power in Pressure Controlled Ventilation (PCV)

**DOI:** 10.1101/2021.07.20.21260873

**Authors:** Christine A. Trinkle, Richard N. Broaddus, Jamie L. Sturgill, Christopher M. Waters, Peter E. Morris

## Abstract

Power is a promising new metric to assess energy transfer from a ventilator to a patient, as it combines the effects of multiple different parameters into a single comprehensive value. For volume-controlled ventilation (VCV), excellent equations exist for calculating power from basic ventilator parameters, but for pressure-controlled ventilation (PCV), an accurate, easy-to-use equation has been elusive. Here, we present a new power equation and evaluate its accuracy compared to the three published PCV power equations. When applied to a sample of 50 patients on PCV with a non-zero rise time, we found that our equation estimated power within an average of 8.4% ± 5.9% (mean ± standard deviation) of the reference value. This new equation is accurate and simple to use, making it an attractive option to estimate power in PCV cases at the bedside.

---

Mechanical power is a promising new metric to evaluate ventilator settings using a single comprehensive value that captures the influence of multiple static and dynamic metrics—PEEP, lung compliance, respiratory rate, and others—resulting in an encompassing picture of energy transfer from a ventilator to the patient. However, many ventilators do not calculate power *in situ. W*hile equations for volume-controlled ventilation (VCV) exist for calculating mechanical power from basic ventilator parameters [1], there remains an opportunity for developing simplified equations for pressure-controlled ventilation (PCV) that can be used at the bedside.

Becher [2] presented two solutions to the PCV power estimation problem. Their “simplified” equation modeled pressure during inspiration as a square wave—removing rise time. As the authors noted, however, this leads to decreased accuracy when rise time is significant. Their “comprehensive” equation accounts for a non-zero rise time, increasing accuracy, but like all complex equations may be challenging for bedside application. Recently, van der Meijden [3] presented a simplified equation, but we and others [4] found that it produces lower accuracy than the comprehensive Becher equation, particularly for patients with long inspiratory time or low flow resistance or compliance (*C*) (Fig. S3).

We developed a new simplified equation by integrating the equation of motion [5] assuming airway pressure (*P*_*aw*_, cmH_2_O) increases linearly from the end-expiratory value (*P*_*PEEP*_, cmH_2_O) to a maximum pressure (*P*_*PEEP*_ + Δ *P*_*insp*_, cmH_2_O) over a prescribed rise time (*t*_*slope*_, s). The result can be represented using a linear model over the entire realistic range of ventilator settings and patient parameters, resulting in a simple, low-error equation for mechanical power (*MP*_*LM*_, Joules/min):

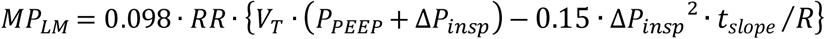

Where *RR* is respiratory rate, *V*_*T*_ is tidal volume (L), Δ *P*_*insp*_ is pressure change from end expiration to end inspiration (cmH_2_O), and *R* is flow resistance (cmH_2_O/L/s). The complete derivation is included in the Supplementary Information.

To evaluate the accuracy of our equation and three published power equations, we used mechanical ventilator data from 50 critically ill patients (Maquet Servo i ventilators, Gothenberg, Sweden) on PCV (Table S1). For each patient, we downloaded ventilator pressure and volume data and integrated the P-V curve numerically to obtain the reference mechanical power value (*MP*_*ref*_).

The equations estimated power within the following mean ± standard deviation of the reference value: our equation, simplified Becher (sB) [2], comprehensive Becher (cB) [2] and van der Meijden (vdM) [3] were 8.4% ± 5.9%, 19.4% ± 12.9%, 10.0% ± 6.8%, and 16.5% ± 14.6% respectively, Fig. 1. The limits of agreement (LoA) for our equation were −1.4 to +5.01 J/min, compared to −5.34 to 12.7 J/min for vdM, - 1.70 to 6.13 J/min for cB, and −3.85 to 12.7 J/min for sB. Additionally, the data used to evaluate all equations includes non-paralyzed patients, while others [2, 3] may have exclusively included patients who were sedated without spontaneous breathing. As demonstrated here, our equation combines the simplicity of the sB and vdM equations with the accuracy of the cB equation, providing an attractive option for bedside calculation of mechanical power.

**Fig. 1.**
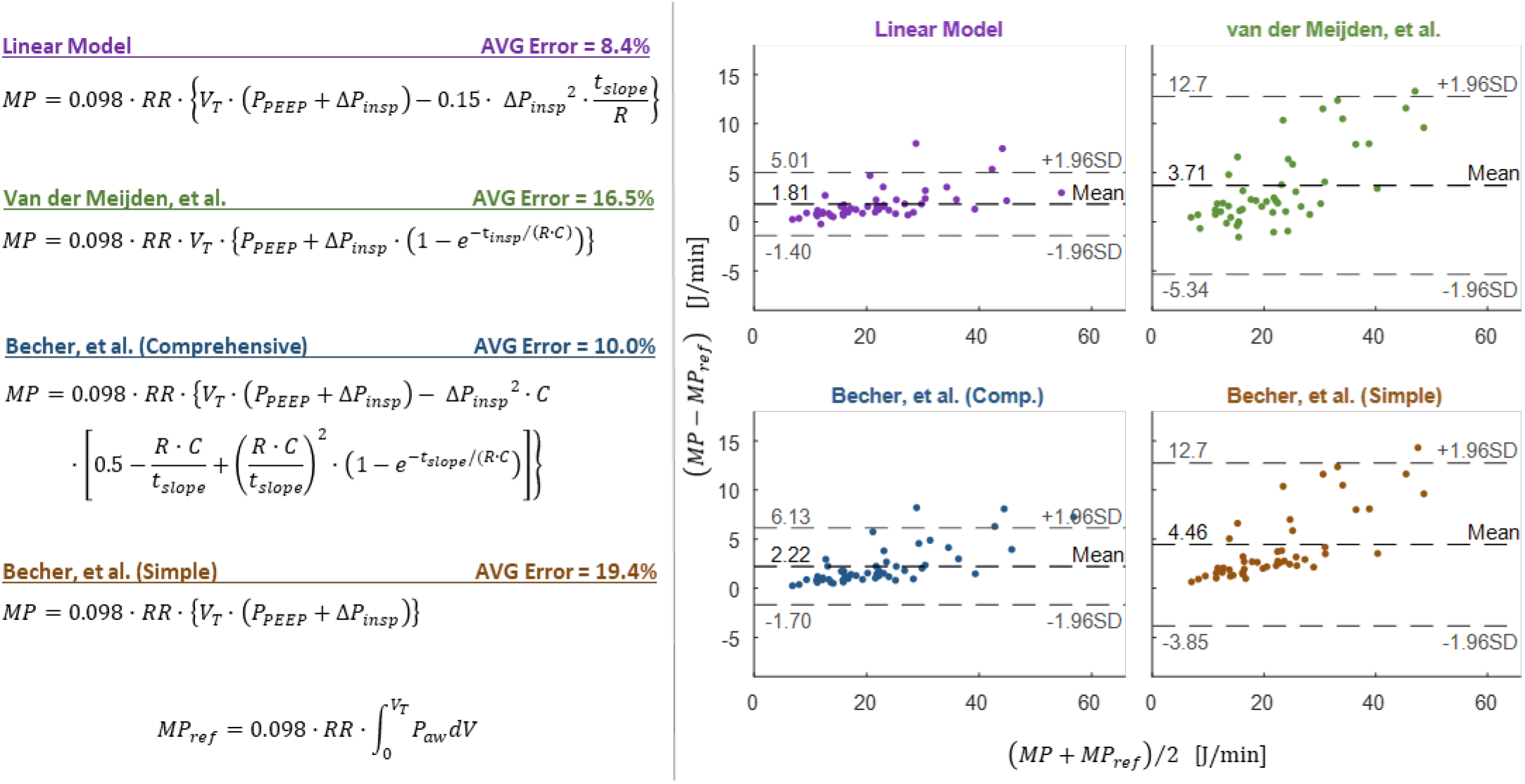
Four equations for estimating power in PCV patients (left), and Bland-Altman plots for each of these equations [2, 3] (right). Bland-Altman plots were generated by plotting the mean of the calculated value (MP) and the reference value (MP_ref_) for each equation (x-axis) against the difference between the calculated value and reference value (y-axis)

## Data Availability

Information about the data set is available upon request.

## Declarations

### Code Availability

not applicable

### Ethics Approval

The study protocol complies with the Declaration of Helsinki and its later amendments and was approved by the University of Kentucky Institutional Review Board and Ethics

### Consent to Participate / Consent to Publish

not applicable

## References

1. Gattinoni, L., T. Tonetti, M. Cressoni, P. Cadringher, P. Herrmann, O. Moerer, A. Protti, M. Gotti, C. Chiurazzi, E. Carlesso, D. Chiumello, and M. Quintel, Ventilator-related causes of lung injury: the mechanical power. Intensive Care Med, 2016. 42(10): p. 1567–1575.

2. Becher, T., M. van der Staay, D. Schädler, I. Frerichs, and N. Weiler, Calculation of mechanical power for pressure-controlled ventilation. Intensive Care Medicine, 2019. 45(9): p. 1321–1323.

3. van der Meijden, S., M. Molenaar, P. Somhorst, and A. Schoe, Calculating mechanical power for pressure-controlled ventilation. Intensive Care Med, 2019. 45(10): p. 1495–1497.

4. Becher, T. and M. van der Staay, Calculation of mechanical power for pressure-controlled ventilation: author’s reply. Intensive Care Med, 2019. 45(10): p. 1498–1499.

5. Marini, J.J. and P.S. Crooke, 3rd, A general mathematicalmodelfor respiratory dynamics relevant to the clinical setting. Am Rev Respir Dis, 1993. 147(1): p. 14–24.

