## Supplementary Information for "Simple, accurate calculation of mechanical power in Pressure Controlled Ventilation (PCV)"

Peter E. Morris [2]

[1] Department of Mechanical Engineering, College of Engineering, University of Kentucky, Lexington, KY, USA

[2] Division of Pulmonary, Critical Care and Sleep Medicine, College of Medicine, University of Kentucky, Lexington, KY, USA

[3] Department of Physiology, College of Medicine, University of Kentucky, Lexington, KY, USA

[4] Saha Cardiovascular Research Center, University of Kentucky, Lexington, KY, USA

### Derivation of Mechanical Power Equation

Mechanical power ($MP$) can be calculated as the integral of the airway pressure ($P_{aw}$) over volume ($V$) during inspiration, or:

$$MP = \int_0^{V_T} P_{aw} dV = \int_0^{t_{insp}} P_{aw} \cdot \frac{dV}{dt} dt \tag{S1}$$

Where $V_T$ represents the tidal volume and $t_{insp}$ is the total inspiratory time. For a simple approximation of mechanical power during pressure controlled ventilation, the airway pressure can be assumed to have a constant value of $P_{aw} = P_{PEEP} + \Delta P_{insp}$ during inspiration, where $P_{PEEP}$ is the positive end expiratory pressure and $\Delta P_{insp}$ is the change in pressure from the end of expiration to the end of inspiration. This results in the "simple" power formula reported by Becher [2]. This can be a reasonable estimation, but incurs error in many cases—particularly when rise time at the beginning of inspiration is non-zero.

To account for non-zero rise time, we assume that the driving pressure waveform takes the same form as assumed by Becher [2] in the derivation of their comprehensive power equation. Briefly, this assumes that at the beginning of each inspiration the airway pressure increases linearly from $P_{aw} = P_{PEEP}$ at time $t = 0$ to $P_{aw} = P_{PEEP} + \Delta P_{insp}$ at time $t = t_{slope}$:

$$P_{aw} = \begin{cases} P_{PEEP} + \Delta P_{insp} \cdot \dfrac{t}{t_{slope}} & when\ 0 \le t < t_{slope} \\ P_{PEEP} + \Delta P_{insp} & when\ t \ge t_{slope} \end{cases} \tag{S2}$$

Using this assumption, it is possible to derive the "comprehensive" mechanical power equation presented by Becher [2]:

$$MP = 0.098 \cdot RR \cdot \left\{ V_T \cdot \left(P_{PEEP} + \Delta P_{insp}\right) - \Delta P_{insp}{}^2 \cdot C \cdot \left[ 0.5 - \frac{R \cdot C}{t_{slope}} + \left(\frac{R \cdot C}{t_{slope}}\right)^2 \cdot \left(1 - e^{-t_{slope}/(R \cdot C)}\right) \right] \right\} \tag{S3}$$

In order to account for the most common units of these parameters in clinical practice, this equation includes a conversion factor of 0.098 Pa*m$^3$/(L*cm$H_2O$). It assumes volumes in Liters, pressures in cm$H_2O$, respiratory rate in breaths per minute, time in seconds, compliance ($C$) in L/cm$H_2O$, and flow resistance ($R$) in cm$H_2O$/L/sec.

While this equation is more accurate than the simple equation, it is complex, and therefore challenging to implement in practice—particularly at the bedside. To simplify this equation, it is possible to derive a linear approximation of equation (S3) for all reasonable values of ventilator and patient parameters. To do so, first we define the term in the square brackets in equation (S3) as:

$$f(t_{slope}/(R \cdot C)) = 0.5 - \frac{R \cdot C}{t_{slope}} + \left(\frac{R \cdot C}{t_{slope}}\right)^2 \cdot \left(1 - e^{-t_{slope}/(R \cdot C)}\right) \tag{S4}$$

Or, making the substitution $x = t_{slope}/(R \cdot C)$, this becomes:

$$f(x) = 0.5 - \frac{1}{x} + \frac{1}{x^2} \cdot (1 - e^{-x}) \tag{S5}$$

For a defined range of $x$ values, $f(x)$ can be approximated by a linear function, $g(x)$. If the range over which the linear approximation is made is $x_1 \le x \le x_2$, one reasonable linear function can be defined as follows:

$$g(x) = \frac{f(x_2) - f(x_1)}{x_2 - x_1} \cdot (x - x_1) + f(x_1) \tag{S6}$$

For lung ventilation, a reasonable lower limit is $x_1 = 0$, which corresponds to a inspiratory rise time of zero ($t_{slope} = 0$). A reasonable upper limit is $x_2 = 0.5$, which can be achieved in a number of ways, including $t_{slope} = 150\ msec$, $R = 10\ cmH_2O/L/sec$, and $C = 0.03\ L/cmH_2O$. Decreasing the value of $t_{slope}$ or increasing the values of $R$ or $C$ would all have the same effect of decreasing $x$, thus we believe that $0 \le x \le 0.5$ represents a reasonable operating range for $x$. And in practice, the equation holds for a range larger than this with minimal error. Therefore, we can approximate $f(x)$ using the following:

$$g(x) = \frac{f(0.5) - f(0)}{0.5 - 0} \cdot (x - 0) + f(0.5) \tag{S7}$$

$$g(x) \approx 0.15 \cdot x \tag{S8}$$

Fig. S1 plots equations (S5) and (S8) over a range of $x$ values, showing good agreement between the original function ($f(x)$) and the linear approximation ($g(x)$).

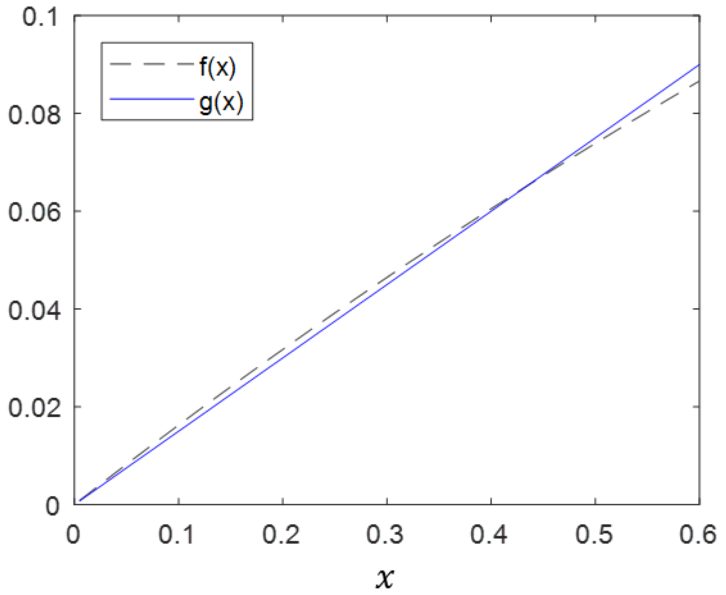


**Fig. S1** Comparison of original function, f(x), and linear approximation, g(x). Functions show excellent agreement over a realistic range of x values

Substituting equation (S8) into equation (S3) in place of $f(x)$ yields:

$$MP = 0.098 \cdot RR \cdot \{V_T \cdot (P_{PEEP} + \Delta P_{insp}) - \Delta P_{insp}{}^2 \cdot C \cdot [0.15 \cdot x]\} \quad \text{(S9)}$$

Once again utilizing the substitution $x = t_{slope}/(R \cdot C)$ and making some minor rearrangements yields the final form of the linear model:

$$\boldsymbol{MP_{LM} = 0.098 \cdot RR \cdot \{V_T \cdot (P_{PEEP} + \Delta P_{insp}) - 0.15 \cdot \Delta P_{insp}{}^2 \cdot t_{slope}/R\}} \quad \textbf{(S10)}$$

Fig. S2 shows the mechanical power predicted by the original comprehensive Becher power equation compared to the power predicted by the linear model derived here. For this figure, $V_T = 0.5\ L, \Delta P_{insp} = 20\ cmH_2O$, $P_{PEEP} = 8\ cmH_2O$, $C = 0.03\ L/cmH_2O$, and $RR = 20\ breaths/min$.

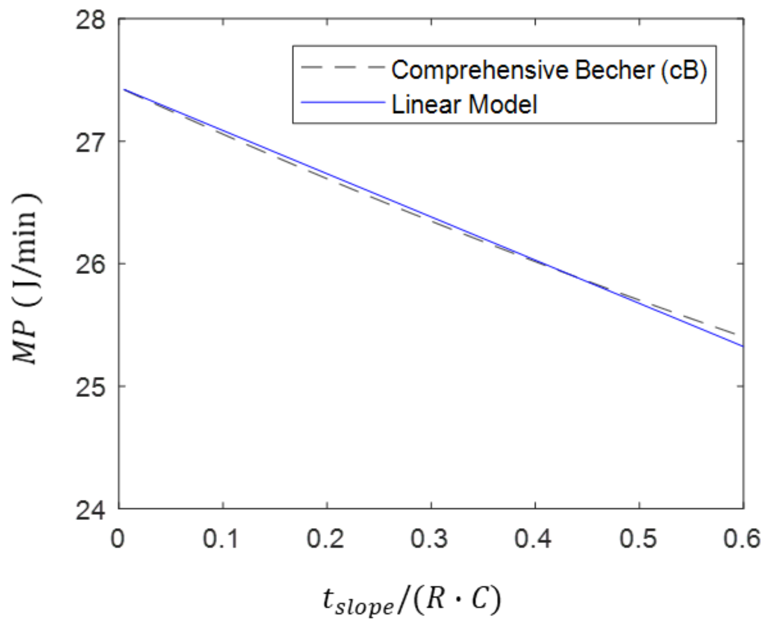


**Fig. S2** Comparison of mechanical power predicted by the comprehensive Becher, et al. equation and the linear model derived here. The two equations show excellent agreement over a realistic range of $t_{slope}/(RC)$ values

### Supplemental Comparison Image

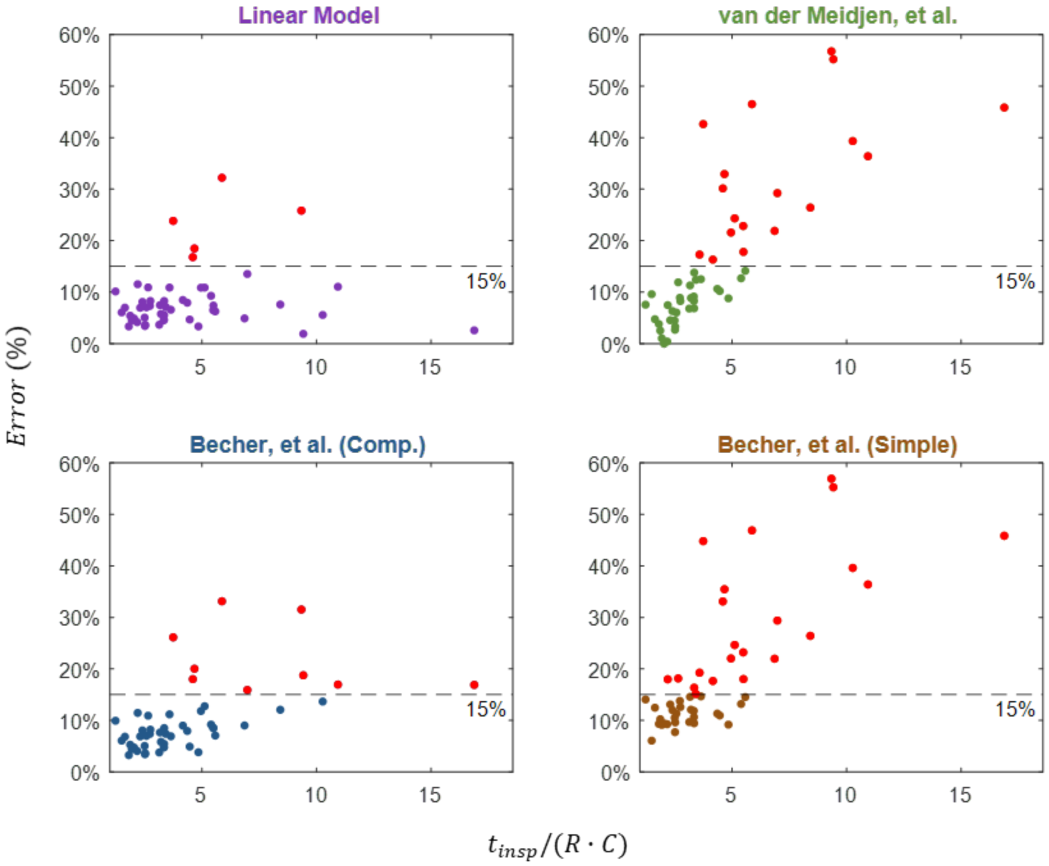


**Fig. S3** Calculation error $(MP_{ref} - MP)/MP_{ref}$ for each of the four equations [2, 3] as a function of the parameter $t_{insp}/(R{\cdot}C)$. Calculations that resulted in greater than 15% error are highlighted in red

**Patient Data Summary**

**Table S1** Selected patient and ventilator parameters (N = 50 subjects)

| Patient Characteristics | Mean Value (Interquartile Range) |
|---|---|
| Age (Years) | 59.4 (24.0 – 84.0) |
| Sex male | 28 (56%) |
| BMI ($kg/m^2$) | 32.8 (16.5 – 66) |
| IBW (kg) | 63.3 (43.1 – 80.0) |
| Total vent days | 16.4 (2.0 – 61.0) |
| P/F ratio | 210.6 (81.0 – 680.0) |
| COPD | 7 (14%) |
| Diabetes | 19 (38%) |
| Cancer | 8 (16%) |
| COVID-19 | 17 (34%) |
| ARDS | 27 (59%) |
| Pneumonia | 26 (52%) |
| Shock | 27 (54%) |
| Paralytic Use | 1 (2%) |
| Patient spontaneously breathing | 21 (42%) |

| Ventilator Characteristics | Mean Value (Interquartile Range) |
|---|---|
| $V_T$/IBW (mL/kg) | 7.622 (5.2 – 10.9) |
| $V_{Min}$ (L/min) | 10.4 (4.8 – 17.4) |
| Respiratory Rate | 22 (14.0 – 36.0) |
| Driving Pressure | 14.4 (5.0 – 29.0) |
| PEEP ($cmH_2O$) | 7.7 (5.0 – 18.0) |
| $C_{dyn}$ ($mL/cmH_20$) | 38.7 (16.0 – 137.0) |
| Flow | 32.3 (21.2 – 59.3) |
| Inspiratory Time | 0.9 (0.6 – 1.2) |
